# Evaluating the potential cost-effectiveness of integrating contraceptive microarray patches into the contraceptive product mix in three low- and middle-income countries

**DOI:** 10.1101/2025.11.05.25339487

**Authors:** Teddy Naddumba, Elisabeth Vodicka, Courtney Jarrahian, Collrane Frivold, Maggie Kilbourne-Brook, Mercy Mvundura

## Abstract

Unmet need for contraceptives still exists within low– and middle-income countries (LMICs). New innovations under development such as the hormonal contraceptive microarray patch (MAP) hold the potential to address some of the reasons for the unmet need, but their value proposition has not been determined. We conducted an analysis to evaluate the potential cost-effectiveness of adding MAPs to the contraceptive product mix in three LMICs.

The analyses were done for Ghana, Nepal, and Senegal, countries with high levels of unmet need. Our analysis evaluated the impact and costs of MAPs meeting a proportion of unmet need for contraceptives, considering various MAP product attributes such as MAP price (range from US$1.65 to US$4.00), market share, and failure rates, among others. Secondary data were used to inform model inputs and assumptions. One-way and probabilistic sensitivity analyses were conducted on key model inputs. Costs and outcomes were evaluated over a 1-year period with reference year 2020. The analysis was done from the health system and limited society perspectives. The per capita gross domestic product (GDP) was the threshold to evaluate cost-effectiveness and was US$2,177, US$1,139, and US$1,490 for Ghana, Nepal, and Senegal, respectively.

With the contraceptive MAP available, and assuming a six-month duration of effectiveness and that 1% of women with unmet need and 1% of current users switch to the contraceptive MAP, we estimated a reduction in pregnancies that would avert 664, 1,076, and 288 disability-adjusted life years, for Ghana, Nepal, and Senegal, respectively, and with cost savings. Cost-effectiveness is impacted by MAP duration of effectiveness, price, and the pregnancy management costs. These findings suggest that the use of new innovations such as MAPs in LMICs has the potential to be cost saving or cost-effective depending on MAP target product attributes and increased use of contraceptives by those with unmet need.

## Introduction

Despite increased availability of contraceptive methods, unmet need for contraceptives still exists within low– and middle-income countries (LMICs). According to the Demographic and Health Surveys, unmet need for contraceptives is calculated as the percentage of sexually active people of reproductive age (15–49 years) who are at risk of pregnancy and are not using any contraceptive method, but want to postpone pregnancy for the next 2 years or do not want to have any more children [1]. A 2019 report estimated that 923 million women of reproductive age in LMICs want to avoid a pregnancy; however, 218 million (24%) of these women have an unmet need for modern contraceptives, with a higher prevalence of unmet need (43%) among adolescents (ages 15–19) [2].

The ongoing presence of unmet need, despite the broad spectrum of contraceptive methods available, is attributed to various factors. Barriers include contraceptive shortages, access (such as transport costs or long wait times for provider-dependent methods), adherence (particularly for daily action methods, such as pills), user need for discretion, cultural factors (such as religion and ethnicity), and concern about side effects [3,4].

New contraceptive technologies in development, such as the hormonal contraceptive microarray patch (MAP) [5], might be able to address some of the reasons for unmet need for contraceptives. MAPs contain micro-projections preloaded with active progestin-only hormonal contraceptive that is released into the body through the outermost layer of the skin. MAPs are potentially applied by the user (self-administered). Depending on the design, the MAP can be removed from the skin immediately after application or might need to be worn for a few minutes before being removed and discarded. Evidence from preclinical studies suggests that contraceptive MAPs may prevent pregnancy for up to 6 months [6]. Therefore, MAPs hold the potential to increase access to contraceptives and meet some of the unmet contraceptive need, as they are being designed to be easy to use for self-administration and could be provided outside the clinic system with minimal prior training for the self-administering user. The product allows discreet use because the MAP is discarded after drug release into the body, and a longer duration of protection (up to 6 months) could improve acceptability and compliance with the regimen compared with other hormonal methods, such as oral pills and injectables.

User preference studies in India, Nigeria, Uganda, Malawi, The Gambia, and the UK have found that self-administered contraceptive MAPs would potentially be desirable and highlighted users’ preferences for key product attributes such as size, administration locations, site reactions, ease of administration, and longer duration of protection [7,8]. Multiple other MAPs are in the early stages of product development for a varying range of indications and therapies such as influenza, measles-rubella vaccines [9] and antiretroviral drug delivery for HIV [10].

Early-stage value propositions of innovations under development can provide evidence to stakeholders such as technology funders and manufacturers on potential value for money and can help identify product profile attributes and use cases that affect potential cost-effectiveness [11]. Previous modeling analyses have been conducted to evaluate the potential cost-effectiveness of vaccine MAPs; these analyses have shown that MAPs potentially could be a cost-saving or cost-effective option to improve immunization coverage [9,12,13,14]. The potential cost-effectiveness of MAPs for non-vaccine drug use cases such as contraceptives, however, has not been evaluated. Our analysis seeks to evaluate the potential cost-effectiveness of incorporating a hormonal contraceptive MAP in the contraceptive product mix in three LMICs.

## Methodology

We evaluate the potential health and economic impact of adding contraceptive MAPs to the contraceptive product mix in three LMICs, Ghana, Nepal, and Senegal, as illustrative countries. We selected these countries because of prevailing high unmet need for contraceptives, which is estimated at 33.6% [15], 24.7% [16], and 21.7% [17] for Ghana, Nepal, and Senegal, respectively. These rates are greater than the average or within the range of global unmet need: an estimated average of 24% [2] for all LMICs.

## Model description

We used a combined decision tree and Markov model to simulate an annual cohort of women aged 15–49 with either met or unmet need for contraceptives in each country over a 1-year time horizon to estimate pregnancy-related costs and health outcomes. The model (Fig 1) includes only women who are at risk of pregnancy; it excludes women who are currently breastfeeding, pregnant, using permanent contraceptive methods, not sexually active or not fecund. We accounted for demographic characteristics in the model to include population composition, prevalence of contraceptive use, and unmet need in addition to parameters related to the current mix of contraceptive methods available in each country. The Markov cycle comprises of monthly cycles in which the model population either moves from the first course of action to second course of action or remains in the current state of action (continue the same contraceptive or no contraceptive method). The model outcomes states are either pregnant not with the pregnancy happening at the event of method failure and later at the end of the year.

**Figure 1.**
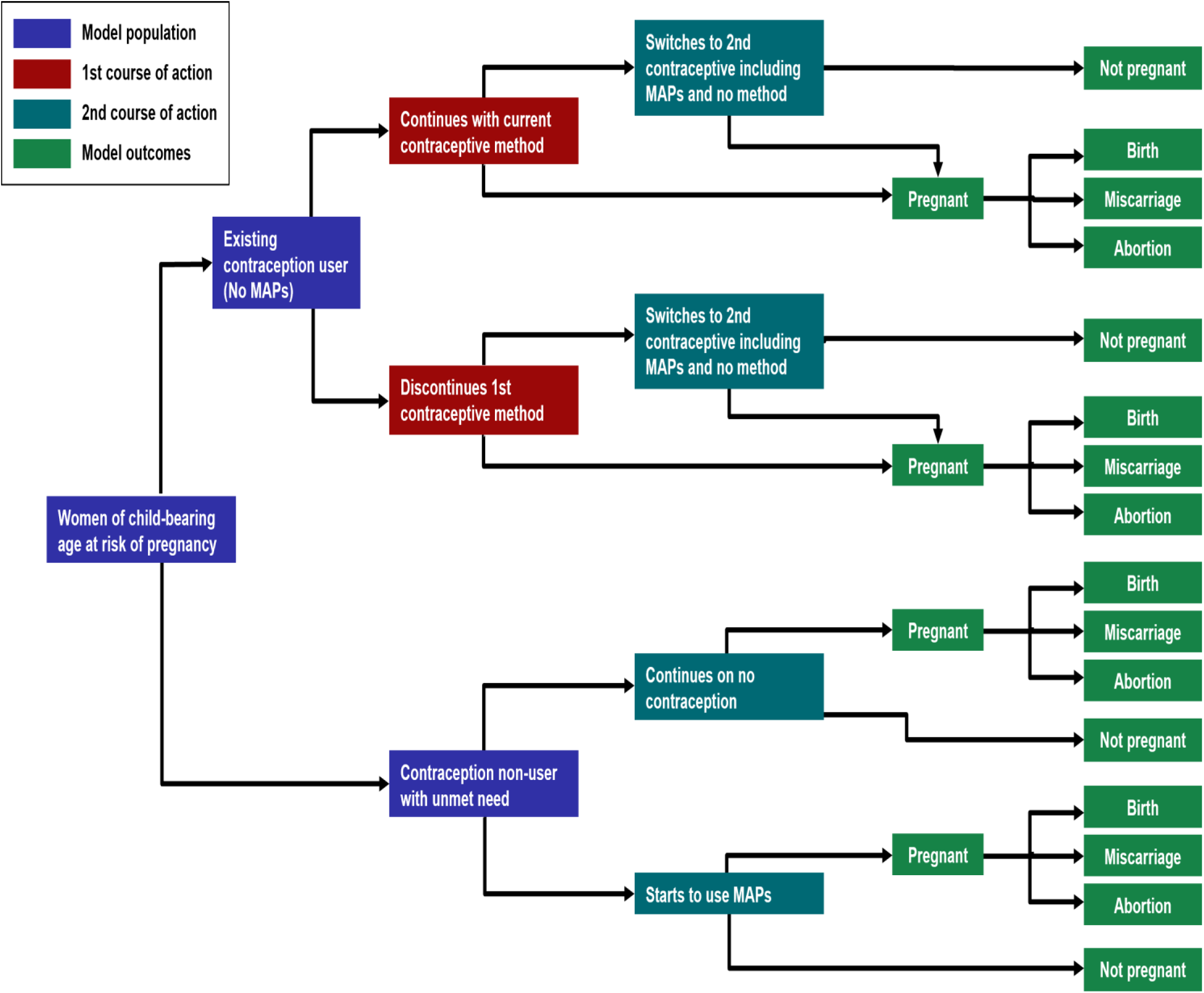
Conceptual framework for decision model evaluating potential health and economic impact of adding a contraceptive MAP to the current method mix in LMICs. Note: The model evaluates a cohort of women of reproductive age with met contraceptive need (i.e., existing contraceptive user) or unmet need (i.e., wanting to avoid pregnancy but currently not on a contraceptive). Women in the model follow two courses of action to capture the introduction of a MAP in the contraceptive product mix. In the first course of action, the women choose to either start a contraceptive method or not. In the second course of action, the women with unmet need either start using the MAP or remain on no method, whereas current contraceptive users choose to either switch to the MAP, discontinue their current contraceptive method, or remain on the current method. The Markov cycle begins at the first course of action and ends with the women either being pregnant or not pregnant.

The model incorporates contraceptive discontinuation rates, potential switching patterns, and typical-use failure rates by method over monthly time cycles. Resulting pregnancies and associated costs, disability-adjusted life years (DALYs), cost per pregnancy prevented, and cost per DALY averted were estimated comparing two scenarios: 1) where there is no MAP in the contraceptive method mix and 2) where there is a MAP available in the contraceptive method mix. The potential impact of the MAP was evaluated as the incremental cost per DALY averted reflected as the ratio of difference in costs and DALYs between the two scenarios. Population characteristics used as model inputs are shown in Table 1, categorized by age group of the women and by study country [18].

**Table 1.**
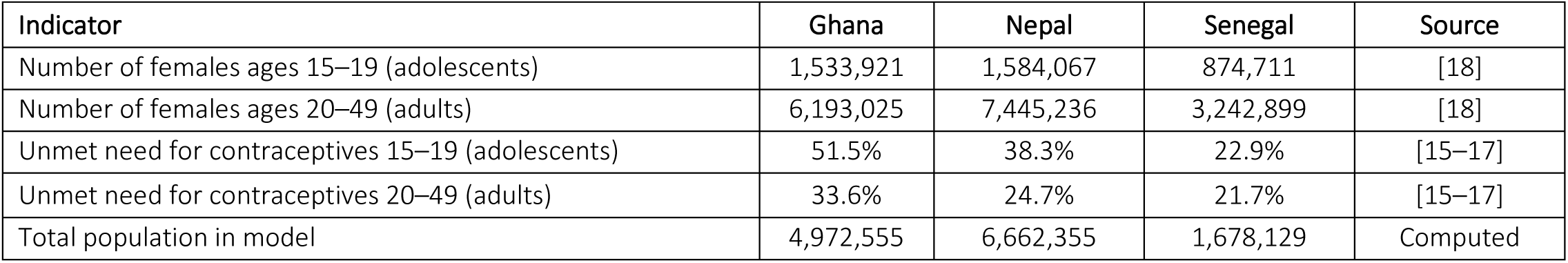
Model population characteristics.

| Indicator | Ghana | Nepal | Senegal | Source |
| --- | --- | --- | --- | --- |
| Number of females ages 15–19 (adolescents) | 1,533,921 | 1,584,067 | 874,711 | [18] |
| Number of females ages 20–49 (adults) | 6,193,025 | 7,445,236 | 3,242,899 | [18] |
| Unmet need for contraceptives 15–19 (adolescents) | 51.5% | 38.3% | 22.9% | [15–17] |
| Unmet need for contraceptives 20–49 (adults) | 33.6% | 24.7% | 21.7% | [15–17] |
| Total population in model | 4,972,555 | 6,662,355 | 1,678,129 | Computed |

Table 2 presents the contraceptive use (uptake) rates by country [15–17], the discontinuation rates [19] of each currently available method, and projected rates for a MAP. Rates for currently available methods reflect estimates from the literature. The uptake of the MAP was explored through varying levels of potential market share for the MAP among non-users and current users of contraceptives who switched to MAPs. Findings from a study of women’s contraceptive discontinuation and switching behavior in Senegal were the source of the rate of transition from first choice of contraceptive to second action of either switching to a new method (existing or future MAP) or discontinuing contraceptive use. These rates were applied to all three countries due to limited data from Ghana and Nepal. Contraceptive failure rates were taken from a multiregional study of contraceptive failure with typical use in 43 countries [20]. For the MAP parameters, inputs such as discontinuation and failure rates were assumed to be similar to injectable contraceptives, given the similarity in hormonal compositions.

**Table 2.**
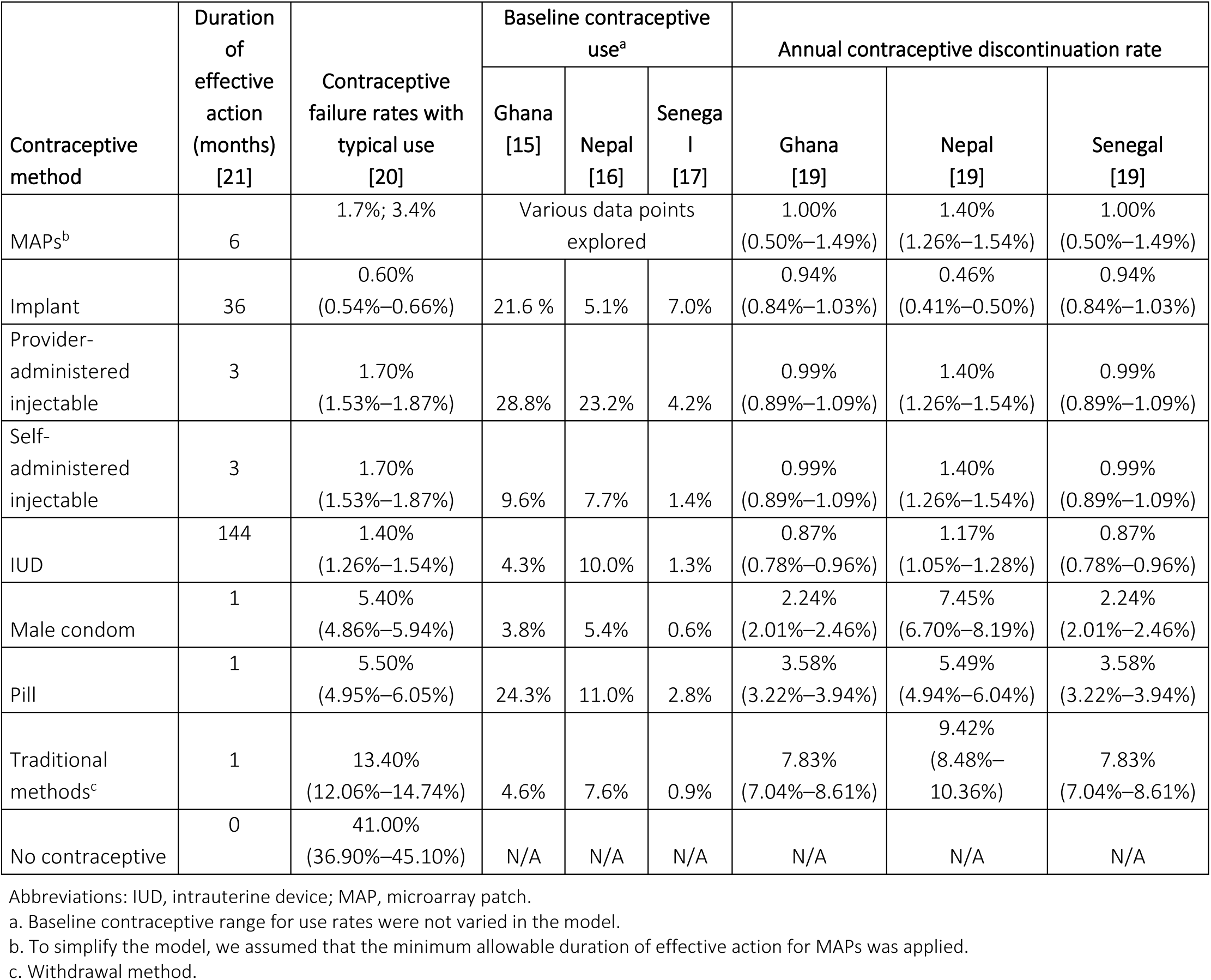
Contraceptive use, efficacy, and probability of discontinuation after 12 months of use by contraceptive method.

| Contraceptive method | Duration of effective action (months) [21] | Contraceptive failure rates with typical use [20] | Baseline contraceptive use<sup>a</sup> |  |  | Annual contraceptive discontinuation rate |  |  |
| --- | --- | --- | --- | --- | --- | --- | --- | --- |
|  |  |  | Ghana [15] | Nepal [16] | Senegal [17] | Ghana [19] | Nepal [19] | Senegal [19] |
| MAPs<sup>b</sup> | 6 | 1.7%; 3.4% | Various data points explored |  |  | 1.00%<br>(0.50%–1.49%) | 1.40%<br>(1.26%–1.54%) | 1.00%<br>(0.50%–1.49%) |
| Implant | 36 | 0.60%<br>(0.54%–0.66%) | 21.6 % | 5.1% | 7.0% | 0.94%<br>(0.84%–1.03%) | 0.46%<br>(0.41%–0.50%) | 0.94%<br>(0.84%–1.03%) |
| Provider-administered injectable | 3 | 1.70%<br>(1.53%–1.87%) | 28.8% | 23.2% | 4.2% | 0.99%<br>(0.89%–1.09%) | 1.40%<br>(1.26%–1.54%) | 0.99%<br>(0.89%–1.09%) |
| Self-administered injectable | 3 | 1.70%<br>(1.53%–1.87%) | 9.6% | 7.7% | 1.4% | 0.99%<br>(0.89%–1.09%) | 1.40%<br>(1.26%–1.54%) | 0.99%<br>(0.89%–1.09%) |
| IUD | 144 | 1.40%<br>(1.26%–1.54%) | 4.3% | 10.0% | 1.3% | 0.87%<br>(0.78%–0.96%) | 1.17%<br>(1.05%–1.28%) | 0.87%<br>(0.78%–0.96%) |
| Male condom | 1 | 5.40%<br>(4.86%–5.94%) | 3.8% | 5.4% | 0.6% | 2.24%<br>(2.01%–2.46%) | 7.45%<br>(6.70%–8.19%) | 2.24%<br>(2.01%–2.46%) |
| Pill | 1 | 5.50%<br>(4.95%–6.05%) | 24.3% | 11.0% | 2.8% | 3.58%<br>(3.22%–3.94%) | 5.49%<br>(4.94%–6.04%) | 3.58%<br>(3.22%–3.94%) |
| Traditional methods<sup>c</sup> | 1 | 13.40%<br>(12.06%–14.74%) | 4.6% | 7.6% | 0.9% | 7.83%<br>(7.04%–8.61%) | 9.42%<br>(8.48%–10.36%) | 7.83%<br>(7.04%–8.61%) |
| No contraceptive | 0 | 41.00%<br>(36.90%–45.10%) | N/A | N/A | N/A | N/A | N/A | N/A |

Abbreviations: IUD, intrauterine device; MAP, microarray patch.
a. Baseline contraceptive range for use rates were not varied in the model.
b. To simplify the model, we assumed that the minimum allowable duration of effective action for MAPs was applied.
c. Withdrawal method.

### Model assumptions

Transitions from the first course of action to the second within the model time horizon, as shown in Fig 1, were explored based on various assumptions. The model assumed that only one pregnancy episode could happen per woman within the horizon of 1 year. Also, while we accounted for potential product switches, for simplicity we assumed that women could switch their contraceptive method only once during the year and would not reinitiate the same contraceptive method after discontinuing it during that same year. The Markov model applied cycle lengths of 1 month; therefore, any product switches and resulting effects were calculated for each month over the 1-year time frame of the model.

We modeled the minimum period of effective action for short-acting methods, such as oral contraceptives, condoms, and traditional methods, as 1 month. For long-term reversible methods such as intrauterine devices (IUDs) or implants, we assumed a 1-month period of effective action because these devices can be removed before the end of the duration of effective action, and fertility is regained after removal. Because of the monthly model cycles, women who were modeled to remain on the same long-acting product for the duration of the model time frame (i.e., did not discontinue for at least 12 months) would in effect experience a cumulative duration of effective action of 12 months due to the long-acting nature of the product. As the duration of effective action of a future MAP product is unknown, we modeled both 3-month and 6-month scenarios with the assumption that the MAP could not be discontinued until the end of that 3-or 6-month time period. Variations in product attributes were assessed in a sensitivity analysis.

### Health outcomes

The health impact of the MAP was calculated as the difference in the estimated number of pregnancies in the scenario with MAPs versus the baseline with no MAPs. Pregnancy outcomes were estimated using the assumptions in Table 3, which shows the probability of abortions, miscarriages, and live births for each studied country. We estimate DALYs per pregnancy outcome using data on DALYs for maternal conditions and complications from the Global Burden of Disease study [22]. DALYs for pregnancy-related conditions are predominately composed of pregnancy and maternal complications such as pre-eclampsia, sepsis and hemorrhages and deaths, rather than morbidity.

**Table 3.** Pregnancy outcomes and associated DALYs per outcome.

| Pregnancy outcomes | Probability of outcome (± 5% sensitivity) | | | DALYs per outcome (± 10% sensitivity) | | |
| --- | --- | --- | --- | --- | --- | --- |
|  | Ghana [23] | Nepal [24] | Senegal [25] | Ghana [22] | Nepal [22] | Senegal [22] |
| Births | 78.00%<br>(74%–82%) | 82.00%<br>(78%–86%) | 76.00%<br>(72%–80%) | 0.105<br>(0.094–0.115) | 0.179<br>(0.161–0.196) | 0.119<br>(0.107–0.131) |
| Miscarriages | 12.00%<br>(11%–13%) | 9.00%<br>(8.55%–9.45%) | 16.00%<br>(15.20%–16.80%) | 0.017<br>(0.015–0.018) | 0.023<br>(0.021–0.025) | 0.118<br>(0.106–0.130) |
| Abortions | 10.00%<br>(9.50%–10.50%) | 9.00%<br>(8.55%–9.45%) | 8.00%<br>(7.60%–8.40%) | 0.017<br>(0.015–0.018) | 0.023<br>(0.021–0.025) | 0.118<br>(0.106–0.130) |

Abbreviation: DALY, disability-adjusted life year.

### Cost inputs

The analysis was done from both the health system and limited societal perspectives. The time horizon was 1 year and hence no discounting was done. All costs were valued in United States dollars (US$) and adjusted to 2020 dollars using the World Bank gross domestic product (GDP) deflator [26].

Costs per contraceptive method are shown for a period aligned with the duration of effective action and include product procurement and service delivery costs (Table 4). Product costs were obtained from United Nations Population Fund (UNFPA) data [21]. The product attributes of MAPS are unknown because they are under development; thus, we explored prices of US$1.60, US$2.40, US$3.20, and US$4.00 per MAP, the lowest MAP price at double the price of injectable contraceptives.

**Table 4.** Input parameters for contraceptive service delivery and access costs.

| | Product costs per period of effective action | Contraceptive service delivery costs per period of effective action (sensitivity ± 50%) | | | Client costs for accessing contraception per visit (sensitivity ± 50%) | | |
| --- | --- | --- | --- | --- | --- | --- | --- |
| Cost inputs | Unit cost [21] | Ghana [28] | Nepal [31] | Senegal [28] | Ghana [28] | Nepal [29] | Senegal [28] |
| MAP<sup>a</sup> | \$1.60, \$2.40, \$3.20, \$4.00 | \$0.46<br>(\$0.23–\$0.68) | \$0.40<br>(\$0.20–0.60) | \$0.28<br>(\$0.14–\$0.42) | \$0.84<br>(\$0.42–\$1.25) | \$2.48<br>(\$1.24–\$3.71) | \$0.76<br>(\$0.38–\$1.14) |
| Implant | \$8.50 | \$4.98<br>(\$2.49–\$7.47) | \$5.94<br>(\$2.97–\$8.91) | \$6.44<br>(\$3.22–\$9.65) | \$0.84<br>(\$0.42–\$1.25) | \$2.48<br>(\$1.24–\$3.71) | \$0.76<br>(\$0.38–\$1.14) |
| Provider-administered injectable | \$0.80 | \$0.90<br>(\$0.45–\$1.35) | \$0.80<br>(\$0.40–\$1.19) | \$0.56<br>(\$0.28–\$0.84) | \$0.84<br>(\$0.42–\$1.25) | \$2.48<br>(\$1.24–\$3.71) | \$0.76<br>(\$0.38–\$1.14) |
| Self-administered injectable | \$0.85 | \$0.62<br>(\$0.31–\$0.92) | \$0.59<br>(\$0.29–\$0.88) | \$0.57<br>(\$0.29–\$0.86) | \$0.21<br>(\$0.10–\$0.31) | \$0.62<br>(\$0.31–\$0.93) | \$0.19<br>(\$0.09–\$0.28) |
| IUD | \$0.44 | \$2.78<br>(\$1.39–\$4.17) | \$3.20<br>(\$1.60–\$4.80) | \$3.16<br>(\$1.58–\$4.74) | \$0.84<br>(\$0.42–\$1.25) | \$2.48<br>(\$1.24–\$3.71) | \$0.76<br>(\$0.38–\$1.14) |
| Male condom | \$0.32 | \$0<br>(\$0–\$0) | \$0<br>(\$0–\$0) | \$0<br>(\$0–\$0) | \$0<br>(\$0–\$0) | \$0<br>(\$0–\$0) | \$0<br>(\$0–\$0) |
| Pill | \$0.18 | \$0.19<br>(\$0.09–\$0.28) | \$0.20<br>(\$0.10–\$0.30) | \$0.59<br>(\$0.29–\$0.88) | \$0.84<br>(\$0.42–\$1.25) | \$2.48<br>(\$1.24–\$3.71) | \$0.76<br>(\$0.38–\$1.14) |
| Traditional method | \$0 | \$0<br>(\$0–\$0) | \$0<br>(\$0–\$0) | \$0<br>(\$0–\$0) | \$0<br>(\$0–\$0) | \$0<br>(\$0–\$0) | \$0<br>(\$0–\$0) |
| Pregnancy costs by country and outcome |  | Ghana [30] |  | Nepal [32] |  | Senegal [30] |  |
| | Births | \$242.67<br>(\$121.33–\$364) | | \$100.58<br>(\$50.29–\$150.86) | | \$80.27<br>(\$40.13–\$120.40) | |
| | Miscarriages | \$50.80<br>(\$25.40–\$76.20) | | \$22.13<br>(\$11.06–\$33.19) | | \$15.94<br>(\$7.97–\$23.91) | |
| | Abortions | \$36.12<br>(\$18.06 –\$54.17) | | \$34.42<br>(\$17.21–\$51.62) | | \$11.04<br>(\$5.52–\$16.55) | |

Abbreviations: IUD, intrauterine device; MAP, microarray patch.
<sup>a</sup> For the sensitivity analysis all costs inputs were varied by ± 50% except for the MAP unit price, which was varied at various price points.

Using data from a study that evaluated the costs of administering injectable contraceptives [27], we assumed service delivery costs to include the health worker trainings, human resources time for counselling and administering the provider-based contraceptive methods, supplies and drugs used for treatment of side effects. Contraception-related side effects are minimal. Headaches are the most observed type, and these are treated with painkillers. For MAPs, service delivery costs were assumed to be half the cost of provider-administered injectable contraceptives.

Client costs for accessing contraception services included transportation costs that we set based on published literature [28,29]. Similarly, for pregnancy outcomes, the cost for each outcome was estimated using data from the literature [30]. Births costs included the cost of antenatal care, delivery, and management of other obstetric and newborn complications. Miscarriage costs were assumed to be the cost of emergency prereferral care, whereas abortion costs included the cost of managing any post-abortion complications [30].

### Model outcomes and threshold for evaluating cost-effectiveness

Incremental cost-effectiveness ratios (ICERs) were calculated as the difference in costs divided by the difference in outcomes (DALYs) between the scenario without MAPs and the scenario with MAPs. The ratios generated the incremental cost per DALY averted. The incremental costs per DALY averted were assessed against a willingness-to-pay threshold based on the per capita GDP for each country [33] (US$2,177 in Ghana, US$1,139 in Nepal, and US$1,490 in Senegal [2020 US$] [34]), as these are implicit thresholds of one time the GDP per capita in LMICs.

### Sensitivity analysis

One-way sensitivity analysis was conducted to evaluate the impact of key model inputs on the cost per DALY averted. To further understand the impact of these attributes on cost-effectiveness of the MAP, we evaluated the impact of varying key inputs for the MAP, such as the duration of effective action, product cost, market share for women with unmet need, and annual failure rate. Uncertainty analysis was conducted for all model inputs, excluding demographics and market share. Results are presented in a tornado diagram to illustrate how the key inputs impact the base-case ICER when individually varied across values within their range. A probabilistic sensitivity analysis was conducted over 5,000 Monte Carlo simulations to estimate the effect of combined uncertainty across all model parameters on the estimated results.

## Results

In the absence of a MAP in the contraceptive mix (scenario 1), we estimate that approximately 855,713 pregnancies would occur in Ghana, 822,175 in Nepal, and 268,227 in Senegal. Associated costs and DALYs are presented in Table 5. With the availability of contraceptive MAPs (scenario 2), assuming 1% of women with unmet need and 1% of current users switching to the MAP, we estimate a reduction in pregnancies by 7,768 in Ghana, 7,150 in Nepal, and 2,417 in Senegal, which would avert 664, 1,076, and 288 DALYs, respectively. From a societal perspective, cost savings with MAPs were estimated at US$1,555,353 (Ghana), US$1,033,309 (Nepal), and US$191,017 (Senegal). The analysis suggests that across the three focus countries, from both a societal and health system perspective, the scenario with a contraceptive MAP in the contraceptive product mix is more effective and cost saving than the scenario without a MAP. Cost savings are attributed to reduced service delivery costs and improved health outcomes that would result from averting additional pregnancies when the unmet contraceptive need is addressed via introduction of a MAP contraceptive.

**Table 5.** Results of cost-effectiveness analysis under base-case assumptions^a^.

|  |  | Ghana | Nepal | Senegal |
| --- | --- | --- | --- | --- |
| **Pregnancies: MAP not available** |  | 855,713 | 822,175 | 268,227 |
| **Mean (95% CR<sup>b</sup>)** |  | (606,685–894,718) | (718,717–918,012) | (231,640–306,440) |
| **Pregnancies: MAP available** |  | 847,946 | 815,025 | 265,811 |
| **Mean (95% CR)** |  | (551,773–899,685) | (730,864–930,378) | (234,587–309,554) |
| **Pregnancies averted** |  | 7,768 | 7,150 | 2,417 |
| **Mean (95% CR)** |  | (4,862–8,757) | (6,653–10,471) | (1,492–2,697) |
| **DALYs: MAP not available** |  | 73,165 | 123,694 | 31,947 |
| **Mean (95% CR)** |  | (44,890–78,391) | (86,301–136,834) | (25,875–38,868) |
| **DALYs: MAP available** |  | 72,501 | 122,619 | 31,659 |
| **Mean (95% CR)** |  | (46,357–78,499) | (86,619–139,510) | (26,031–39,086) |
| **DALYs averted** |  | 664 | 1,076 | 288 |
| **Mean (95% CR)** |  | (409–874) | (777–1,583) | (169–336) |
| **Societal perspective** | **Costs: MAP not available** | \$199,769,696 | \$171,955,189 | \$29,641,689 |
| | **Mean (95% CR)** | (\$40,835,974–\$238,081,633) | (\$105,157,916–\$245,509,595) | (\$16,913,631–\$50,016,849) |
| | **Costs: MAP available** | \$198,214,343 | \$170,921,880 | \$29,450,222 |
| | **Mean (95% CR)** | (\$57,884,698–\$240,607,107) | (\$106,474,912–\$248,458,503) | (\$17,275,102–\$50,557,722) |
| | **Cost difference with MAP available** | -\$1,555,353 | -\$1,033,309 | -\$191,017 |
| | **Mean (95% CR)** | (-248,812– -2,917,736) | (-\$109,996– -\$3,028,890) | (-\$58,620– -\$619,138) |
|  | **ICER** | Dominant | Dominant | Dominant |
| | **Costs: MAP not available** | \$191,236,724 | \$121,029,644 | \$27,287,936 |
| | **Mean (95% CR)** | (\$51,936,559–\$231,401,405) | (\$63,152,635–\$188,458,239) | (\$14,944,695–\$48,739,795) |
| | **Costs: MAP available** | \$189,684,357 | \$120,177,809 | \$27,094,767 |
| | **Mean (95% CR)** | (\$50,598,068–\$234,566,168) | (\$61,598,543–\$187,177,522) | (\$15,069,278–\$47,952,555) |
| | **Cost difference with MAP available** | -\$1,552,366 | -\$851,835 | -\$192,862 |
| | **Mean (95% CR)** | (-\$234,740– -\$2,914,736) | (-\$141,955– -\$3,022,887) | (-\$51,446– -\$590,655) |
|  | **ICER** | Dominant | Dominant | Dominant |

Abbreviations: CI, confidence interval; CR, credible range; DALY, disability-adjusted life year; ICER, incremental cost-effectiveness ratio; MAP, microarray patch. Dominant: Scenario with MAP available is more effective and less costly than current scenario without a MAP in the contraceptive product mix.
- The base-case analysis assumes that a MAP contraceptive has a 6-month duration of effective action, US\$1.60 price, and 1.7% failure rate. It further assumes that introduction of a MAP contraceptive into the contraception method mix would address 1% of unmet contraceptive need and 1% of current users switching to a MAP. - Credible ranges of 95% were estimated via the probabilistic sensitivity analysis results.

Across the three countries, potential cost savings are projected for a MAP which has a 6-month duration of effective action and 1% market share at the different procurement prices evaluated at US$1.60, US$2.40, US$3.20, and US$4.00 (Table 6). The MAP scenario ceases to be cost saving when the price is higher than US$17.34 for Ghana, US$9.41 for Nepal, and US$7.33 for Senegal, despite averting more DALYs, however, these scenarios are still cost-effective in comparison with the willingness-to-pay threshold of one time the GDP per capita. With a 6-month duration of effectiveness, the model estimates further cost savings for the contraceptive M AP scenario.

**Table 6.** ICERS for the MAP at varying procurement prices.

| MAP price<br>US\$ | Country | Societal perspective | | | Health system perspective | | |
| --- | --- | --- | --- | --- | --- | --- | --- |
| | | Cost difference<br>with MAP available | ICER, US\$ per DALY<br>averted | | Cost difference with<br>MAP available | ICER, US\$ per DALY<br>averted | |
| 1.60 | Ghana | -1,552,366 | -2,337 | Dominant | -1,555,353 | -2,342 | Dominant |
|  | Nepal | -851,835 | -792 | Dominant | -1,033,309 | -961 | Dominant |
|  | Senegal | -192,862 | -670 | Dominant | -191,017 | -664 | Dominant |
| 2.40 | Ghana | -1,473,335 | -2,218 | Dominant | -1,476,322 | -2,223 | Dominant |
|  | Nepal | -746,055 | -694 | Dominant | -927,529 | -862 | Dominant |
|  | Senegal | -166,191 | -578 | Dominant | -164,345 | -571 | Dominant |
| 3.20 | Ghana | -1,394,304 | -2,099 | Dominant | -1,397,290 | -2,104 | Dominant |
|  | Nepal | -640,276 | -595 | Dominant | -821,750 | -764 | Dominant |
|  | Senegal | -139,519 | -485 | Dominant | -137,674 | -478 | Dominant |
| 4.00 | Ghana | -1,315,273 | -1,980 | Dominant | -1,318,259 | -1,985 | Dominant |
|  | Nepal | -534,496 | -497 | Dominant | -715,970 | -666 | Dominant |
|  | Senegal | -112,848 | -392 | Dominant | -111,003 | -386 | Dominant |

Abbreviations: DALY, disability-adjusted life year; ICER, incremental cost-effectiveness ratio; MAP, microarray patch. Dominant: Scenario with MAP available is more effective and less costly than current scenario without a MAP in the contraceptive product mix.

When we assume a higher MAP price combined with a potentially shorter duration of effective action compared with the base case (i.e., 3 months compared with 6 months in the base case), we find that across all scenarios evaluated, MAPs remain cost saving for Ghana (Table 7). However, for Nepal and Senegal the scenario with MAPs is no longer cost saving but the scenario with MAPs remains cost-effective compared with a no-MAP scenario (Table 7). For example, in Senegal, a MAP with a 3-month duration of effectiveness, meeting 1% of unmet need, and priced at US$4.00 with a failure rate of 1.7% is still highly likely to be cost-effective with an ICER of US$196 per DALY averted (approximately 13% of the Senegal 2020 GDP per capita of US$1,490). MAP product prices and live birth costs were the most influential parameters on the ICER. The tornado diagrams in Fig 2 further illustrate the key inputs that most affect the ICER in each of the selected countries.

**Figure 2.**
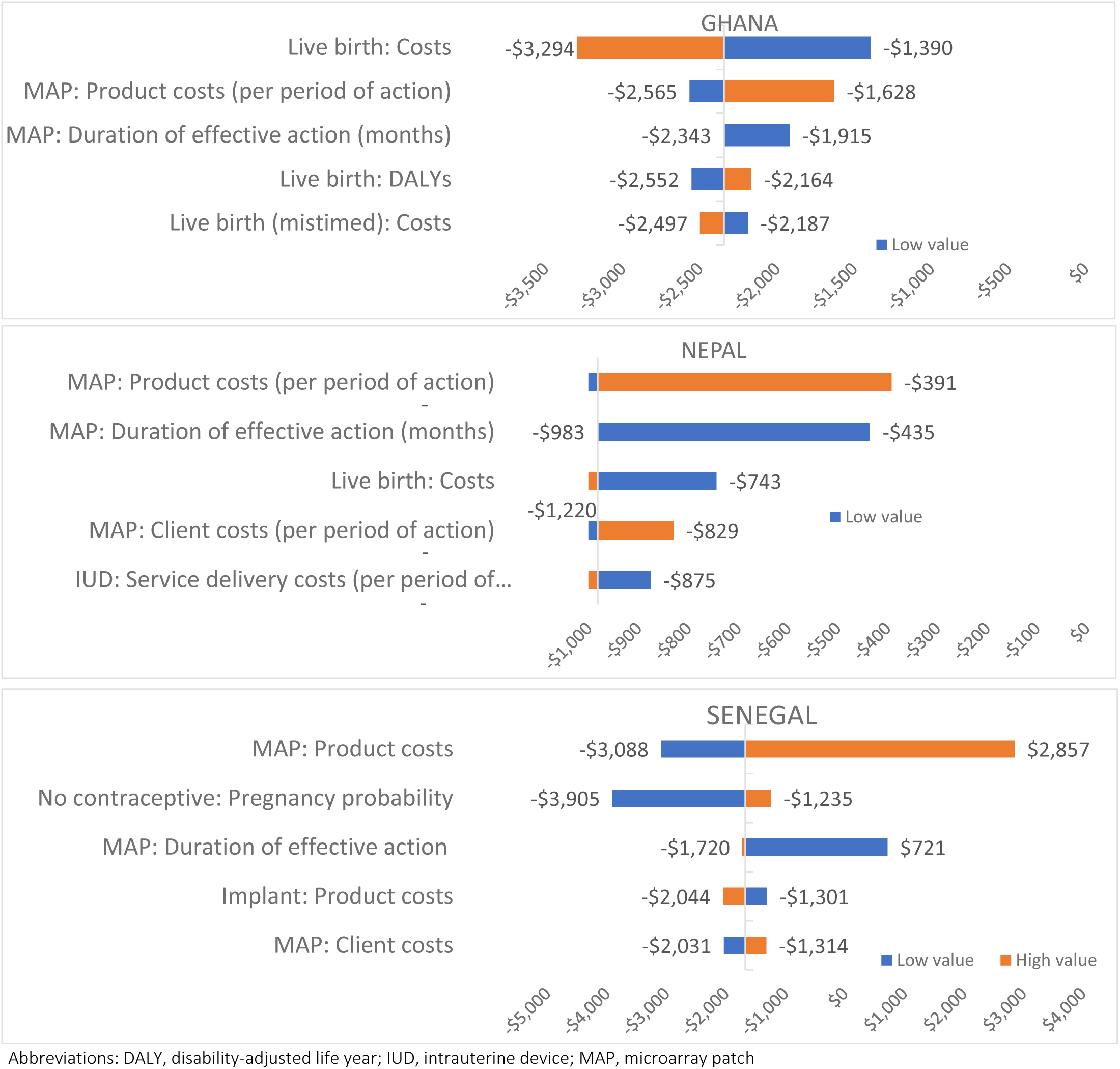
One-way sensitivity analysis for the incremental cost per DALY averted due to variation of key inputs.

**Table 7.** Cost-effectiveness results under varying assumptions for MAP product attributes.

| MAP attributes | Country | Pregnancies:<br>MAP not<br>available | Pregnancies:<br>MAP<br>available | Pregnancies<br>averted | DALYs<br>averted for<br>pregnancies<br>averted | Cost difference<br>with MAP<br>available<br>(MAP<br>procurement<br>price = US\$1.60) | ICER, US\$ per<br>DALY averted<br>(MAP<br>procurement<br>price = US\$1.60) | Cost difference<br>with MAP<br>available<br>(MAP<br>procurement<br>price = US\$2.40) | ICER, US\$ per<br>DALY averted<br>(MAP<br>procurement<br>price = US\$2.40) | Cost difference<br>with MAP<br>available<br>(MAP<br>procurement<br>price = US\$3.20) | ICER, US\$ per<br>DALY averted<br>(MAP<br>procurement<br>price = US\$3.20) | Cost difference<br>with MAP<br>available<br>(MAP<br>procurement<br>price = US\$4.00) | ICER, US\$ per<br>DALY averted<br>(MAP<br>procurement price<br>= US\$4.00) |
| --- | --- | --- | --- | --- | --- | --- | --- | --- | --- | --- | --- | --- | --- |
| 6-month duration of effective action<br>0% market share of unmet need<br>1% of users switch to MAPs<br>1.7% failure rate | Ghana | 855,713 | 855,440 | 274 | 23 | -\$228,790 | Dominant | -\$195,386 | Dominant | -\$161,982 | Dominant | -\$128,578 | Dominant |
| | Nepal | 822,175 | 821,404 | 771 | 116 | -\$692,206 | Dominant | -\$625,257 | Dominant | -\$558,308 | Dominant | -\$491,358 | Dominant |
| | Senegal | 268,227 | 268,171 | 56 | 7 | -\$86,482 | Dominant | -\$74,179 | Dominant | -\$61,875 | Dominant | -\$49,572 | Dominant |
| 6-month duration of effective action<br>0% market share of unmet need<br>3% of users switch to MAPs<br>1.7% failure rate | Ghana | 855,713 | 854,893 | 821 | 70 | -\$686,370 | Dominant | -\$586,158 | Dominant | -\$485,947 | Dominant | -\$385,735 | Dominant |
| | Nepal | 822,175 | 819,862 | 2,312 | 384 | -\$2,076,618 | Dominant | -\$1,875,771 | Dominant | -\$1,674,923 | Dominant | -\$1,474,075 | Dominant |
| | Senegal | 268,227 | 268,059 | 168 | 20 | -\$259,445 | Dominant | -\$222,536 | Dominant | -\$185,626 | Dominant | -\$148,716 | Dominant |
| 6-month duration of effective action<br>1% market share of unmet need<br>1% of users switch to MAPs<br>3.4% failure rate | Ghana | 855,713 | 848,685 | 7,028 | 601 | -\$1,409,350 | Dominant | -\$1,330,644 | Dominant | -\$1,251,938 | Dominant | -\$1,173,232 | Dominant |
| | Nepal | 822,175 | 816,015 | 6,160 | 927 | -\$949,070 | Dominant | -\$843,725 | Dominant | -\$738,381 | Dominant | -\$633,037 | Dominant |
| | Senegal | 268,227 | 266,061 | 2,166 | 258 | -\$175,296 | Dominant | -\$148,735 | Dominant | -\$122,173 | Dominant | -\$95,612 | Dominant |
| 6-month duration of effective action<br>3% market share of unmet need<br>3% of users switch to MAPs<br>3.4% failure rate | Ghana | 855,713 | 834,629 | 21,085 | 1,803 | -\$4,228,049 | Dominant | -\$3,991,932 | Dominant | -\$3,755,814 | Dominant | -\$3,519,697 | Dominant |
| | Nepal | 822,175 | 803,695 | 18,480 | 2,780 | -\$2,847,209 | Dominant | -\$2,531,176 | Dominant | -\$2,215,144 | Dominant | -\$1,899,111 | Dominant |
| | Senegal | 268,227 | 261,728 | 6,499 | 774 | -\$525,889 | Dominant | -\$446,204 | Dominant | -\$366,520 | Dominant | -\$286,836 | Dominant |
| 3-month duration of effective action<br>1% market share of unmet need<br>1% of users switch to MAPs<br>1.7% failure rate | Ghana | 855,713 | 847,958 | 7,756 | 663 | -\$1,269,774 | Dominant | -\$1,112,238 | Dominant | -\$954,701 | Dominant | -\$797,165 | Dominant |
| | Nepal | 822,175 | 815,048 | 7,127 | 1,072 | -\$444,200 | Dominant | -\$233,452 | Dominant | -\$22,703 | Dominant | \$188,045 | \$175.38 |
| | Senegal | 268,227 | 265,815 | 2,412 | 287 | -\$103,194 | Dominant | -\$50,029 | Dominant | \$3,136 | \$10.92 | \$56,301 | \$196.00 |
| 3-month duration of effective action<br>3% market share of unmet need<br>3% of users switch to MAPs<br>3.4% failure rate | Ghana | 855,713 | 834,663 | 21,051 | 1,800 | -\$3,378,832 | Dominant | -\$2,909,146 | Dominant | -\$2,439,459 | Dominant | -\$1,969,773 | Dominant |
| | Nepal | 822,175 | 803,759 | 18,416 | 2,771 | -\$1,094,840 | Dominant | -\$466,501 | Dominant | \$161,838 | \$58.41 | \$790,176 | \$285.20 |
| | Senegal | 267,227 | 261,740 | 6,487 | 773 | -\$264,643 | Dominant | -\$106,135 | Dominant | \$52,374 | \$67.78 | \$210,883 | \$272.92 |

Abbreviations: DALY, disability-adjusted life year; ICER, incremental cost-effectiveness ratio; MAP, microarray patch. Dominant: Scenario with MAP available is more effective and less costly than current scenario without a MAP in the contraceptive product mix.

## Discussion

Our findings suggest that contraceptive MAPs hold the potential to be cost saving and or cost-effective, if key target product and market attributes are met, compared with the current contraceptive mix in the three selected countries. Across the range of our analyses, we project that introducing a contraceptive MAP into the market mix would result in 56–21,085 fewer unintended pregnancies compared with the current mix, resulting in a subsequent effect of 7–1,803 DALYs from pregnancy outcomes averted. Total incremental costs ranged from –US$12,997 (a cost savings) to US$285.

The scenario and threshold analyses identified potential situations in which introducing a MAP into the contraceptive mix would likely not be cost-effective. For example, our threshold analysis found that when MAPs are modeled for a high annual failure rate (at a failure rate greater than 14%), MAPs are projected not to be cost-effective for both Nepal and Senegal, as the ICERs (US$5,461 and US$1,672 per DALY averted, respectively) are greater than the cost-effectiveness threshold of one time the GDP per capita compared with a no-MAP scenario. However, 14% would be a high annual failure rate for a hormonal contraceptive method and is thus unlikely for a MAP. Research aggregating typical-use failure rates across 43 countries suggests a median 12-month failure rate ranging from 0.6% with the implant to 5.5% with oral contraceptives, while traditional methods such as withdrawal and periodic abstinence are associated with higher failure rates of 13.4% and 13.9%, respectively [20].

Although there is limited literature for cost-effectiveness analysis of MAP contraceptive products in LMICs for comparison, other alternative contraceptive products have been assessed in high-income countries. For example, transdermal contraceptive patches were demonstrated to be cost saving compared with oral contraceptives in the United States [35]. Our findings are similar to those of the cost-effectiveness study for self-injecting contraceptives conducted in Burkina Faso, Uganda, and Senegal [27], which shows that new contraceptive technology has the potential to be cost-effective. Relatedly, the findings for the cost-effectiveness analysis of hepatitis B vaccine MAPs in LMICs showed that these MAPs increased access/equity and were cost-effective [9]. This is similar to our findings for the contraceptive MAPs where cost-effectiveness was realized with increased access and unmet need addressed.

## Limitations

Due to data limitations and MAPs not yet being available in contraceptive choices, we were unable to capture the full complexity of contraceptive use patterns in the real world, specifically for situations where contraception users might switch methods more than once in a year. Further, our model estimates relied on average data on use and unmet contraceptive need for each selected country; however, age-disaggregated data would provide more detailed information on whether particular age groups would benefit more from a MAP versus another type of contraceptive.

MAPs are early in the development phase, key products attributes are uncertain, and whether they will increase uptake of contraception is unknown at this time. Therefore, for the base case we conservatively used less favorable values for the key product attributes and explored a wide sensitivity range of model scenarios (see Results) to provide broad-ranging estimates of what effect these variables would have on unmet contraceptive need.

Our analysis centers on a limited societal perspective, which focuses on health outcomes for the contraceptive user. However, we find that more cost savings could be realized with the contraceptive MAP if a broader perspective is explored. The broader perspective would account for possible costs and non-health impacts associated with contraceptive use and pregnancies—including caretaker burden, opportunity cost of lost income, and future health costs or non-health impacts, such as treating long-term complications of pregnancy. We did not account for health impact associated with side effects of contraceptives, these are minimal for the general population and have minimal health impact.

Our study focused on the general population, without considering HIV status since WHO guidance has clarified that persons living with HIV and those at-risk of HIV infection can safely use all methods—including hormonal contraceptives. However contraceptive method choice is influenced by complex range of factors, and we understand that HIV-related stigma can have a negative impact on individuals’ health care seeking behavior. We recommend further exploration of the potential impact of contraceptive MAPs in addressing unmet need for contraception amongst people living with HIV and other key populations to further clarify the value proposition for this new drug delivery system.

## Conclusion

These findings suggest that use of contraceptive MAPs in LMICs has the potential to be cost saving or cost-effective if target product attributes are met and if MAPs enable increased use of contraception by those currently with unmet need. Cost-effectiveness is driven by aversion of pregnancies and pregnancy management costs specifically for settings with large populations or high fertility rates. This analysis supports ongoing research and development of contraceptive MAPs by providing guidance on the relative impact of different product attributes that are likely to influence uptake and acceptability.

## Data Availability

All data has been provided within the manuscript.

## Acknowledgment

We acknowledge Nathaniel Hendrix for his contribution to the study, which was instrumental in the design of the model that was used to conduct the cost-effectiveness analysis.

